# Marked heterogeneity in malaria infection risk in a Malian longitudinal cohort

**DOI:** 10.1101/2024.08.20.24312307

**Authors:** Emily LaVerriere, Zachary M. Johnson, Meg Shieh, Nadege Nziza, Galit Alter, Caroline O. Buckee, Peter D. Crompton, Boubacar Traore, Tuan M. Tran, Daniel E. Neafsey

## Abstract

Variation in malaria infection risk, a product of disease exposure and immunity, is poorly understood. We genotypically profiled over 13,000 blood samples from a six-year longitudinal cohort in Mali to characterize malaria infection dynamics with unprecedented detail. We generated *Plasmodium falciparum* amplicon sequencing data from 464 participants (aged 3 months -25 years) across the six-month 2011 transmission season and profiled a subset of 120 participants across the subsequent five annual transmission seasons. We measured infection risk as the molecular force of infection (molFOI, number of genetically distinct parasites acquired over time). We found that molFOI varied extensively among individuals (0-55 in 2011) but was independent of age and consistent within individuals over multiple seasons. Reported bednet usage was nearly universal. The HbS allele associated with lower molFOI, and functional antibody signatures correlated with both low and high molFOI participants, identifying candidate immune correlates of protection and risk, respectively. The large inter-individual variability in molFOI and consistency of intra-individual infection risk over time remains largely unexplained, but should be considered in clinical trials and implementation of malaria interventions. Factors contributing to heterogeneity in infection risk should be further studied to inform development of future malaria interventions.

## Introduction

Disease risk varies widely among hosts for many communicable diseases^1–3^. Risk of disease is a function of both the risk of infection by the pathogen and individual susceptibility to the development of symptomatic disease once infected. For pathogens that can cause asymptomatic infections, characterization of both symptomatic disease and asymptomatic infection is necessary to characterize the infectious reservoir, disease risk among the infected, and transmission dynamics^4^. Understanding variability in infection risk is important for designing and interpreting clinical trial data and evaluations of other potential interventions; if not accounted for, host variability in disease exposure and/or immunity could confound interpretations of intervention efficacy^1,5^. More importantly, a better understanding of what contributes to the heterogeneity in risk of infection could allow interventions to be targeted towards those at highest risk.

Despite the importance of heterogeneous infection risk, it can be difficult to study for diseases like malaria, where the gradual development of naturally acquired immunity (NAI) reduces risk of symptomatic disease without affecting infection risk, masking heterogeneity in infection incidence^5^. Consequently, an area of high transmission is an ideal setting in which to study infection risk; with high enough transmission, low infection incidence may be interpreted as a consequence of low risk, rather than random chance. However, in high-transmission settings, polyclonal infections resulting from newly incident infections in already-infected people will be more common^6^, preventing estimates of infection incidence through binary infection detection methods such as blood smear microscopy or PCR^7^. To better understand complex infection dynamics, a high resolution methodology is required to detect new infections among already-infected individuals.

In this study, we used Illumina-based amplicon sequencing of four highly polymorphic malaria parasite antigens to provide an unparalleled portrait of polyclonal infection dynamics in a high transmission setting over six years for a large cohort of participants. We sequenced *Plasmodium falciparum* DNA from 13,152 dried blood spot samples from a longitudinal cohort in Kalifabougou, Mali (**Figure 1**)^7^. Kalifabougou is a small, rural community with strong, seasonal malaria transmission during approximately half the year from July to December^8^. Approximately half (n=6,432) of the samples we genotyped are from the 2011 season, and represent 464 participants, ages 3 months -25 years, each of whom provided a finger prick blood sample every two weeks during the six month transmission season. The remaining samples (n=6,720) represent a subset of 120 participants (ages 3 months -8 years at enrollment), studied over the subsequent five annual transmission seasons, during which participants provided a blood sample every month. Participants also provided blood samples at any self-referred clinic visits when malaria was suspected by study clinicians. Prior studies of this cohort, using PCR to detect the first infection of the 2011 transmission season in all participants irrespective of symptoms, found that the majority of participants became infected with *P. falciparum* during a single transmission season, establishing Kalifabougou as an appropriately high transmission setting for evaluating infection risk heterogeneity^7^.

**Figure 1.**
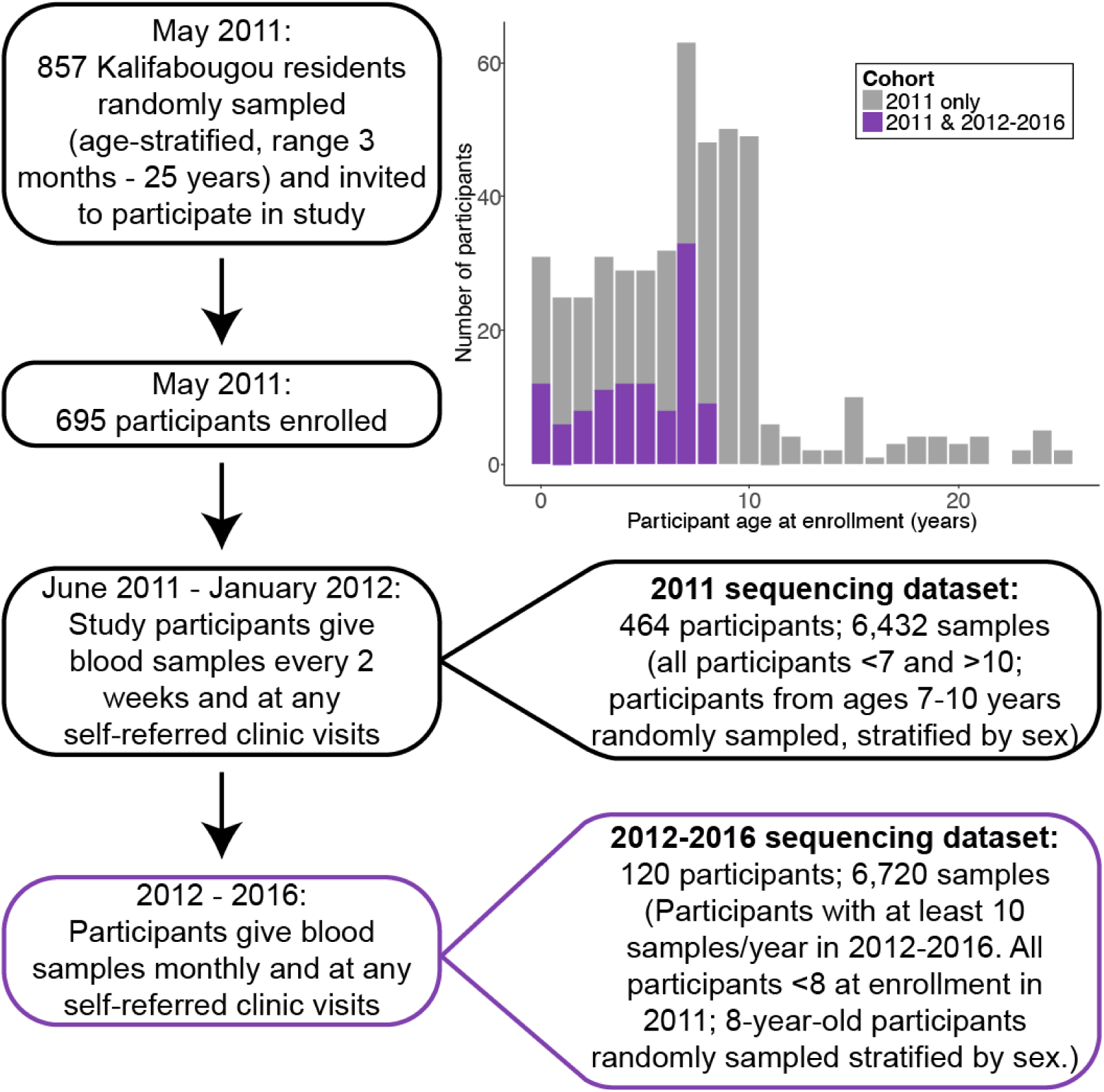
Study participants and timing of sampling within the two sequencing datasets. As described in Tran *et al*., 2013, the Kalifabougou cohort began in May 2011. We sequenced samples from 464 participants from the 2011 transmission season, as well as 120 participants who continued in the study from 2012-2016.

We performed genotyping of participant specimens in order to characterize infection risk heterogeneity and to look for associations between infection risk and features that might drive variation in such risk. We estimated the molecular force of infection (molFOI, the number of genetically distinct strains that infect a participant over a set period of time)^9,10^ (**Extended Data Figure 1)** to summarize infection history in 464 participants across one or more transmission seasons. We explore diverse factors that may contribute to the consistent patterns of heterogeneous infection risk that we observe among participants, including hemoglobin types, demography, geospatial data^11^, and immunological features^12^.

## Results

### molFOI is not associated with age

We define the molecular force of infection (molFOI) as the number of genetically distinct strains that infected a given participant over a period of time, where a newly incident infection by a genetically distinct strain was defined as observation of a novel amplicon haplotype (not seen in the previous 2 samples from the participant) at one or more of the four genotyped loci^10^. We calculated the molFOI for each participant over the entirety of the 2011 season (**Figure 2a**). The molFOI range was 0-55 (mean: 11, median: 10, standard deviation: 9). We found no correlation between participant age and molFOI (quasi-Poisson regression, p-value = 0.077), suggesting NAI that accumulates with age in this population^7^ reduces the risk of clinical disease but does not provide measurable protection against infection.

**Figure 2.**
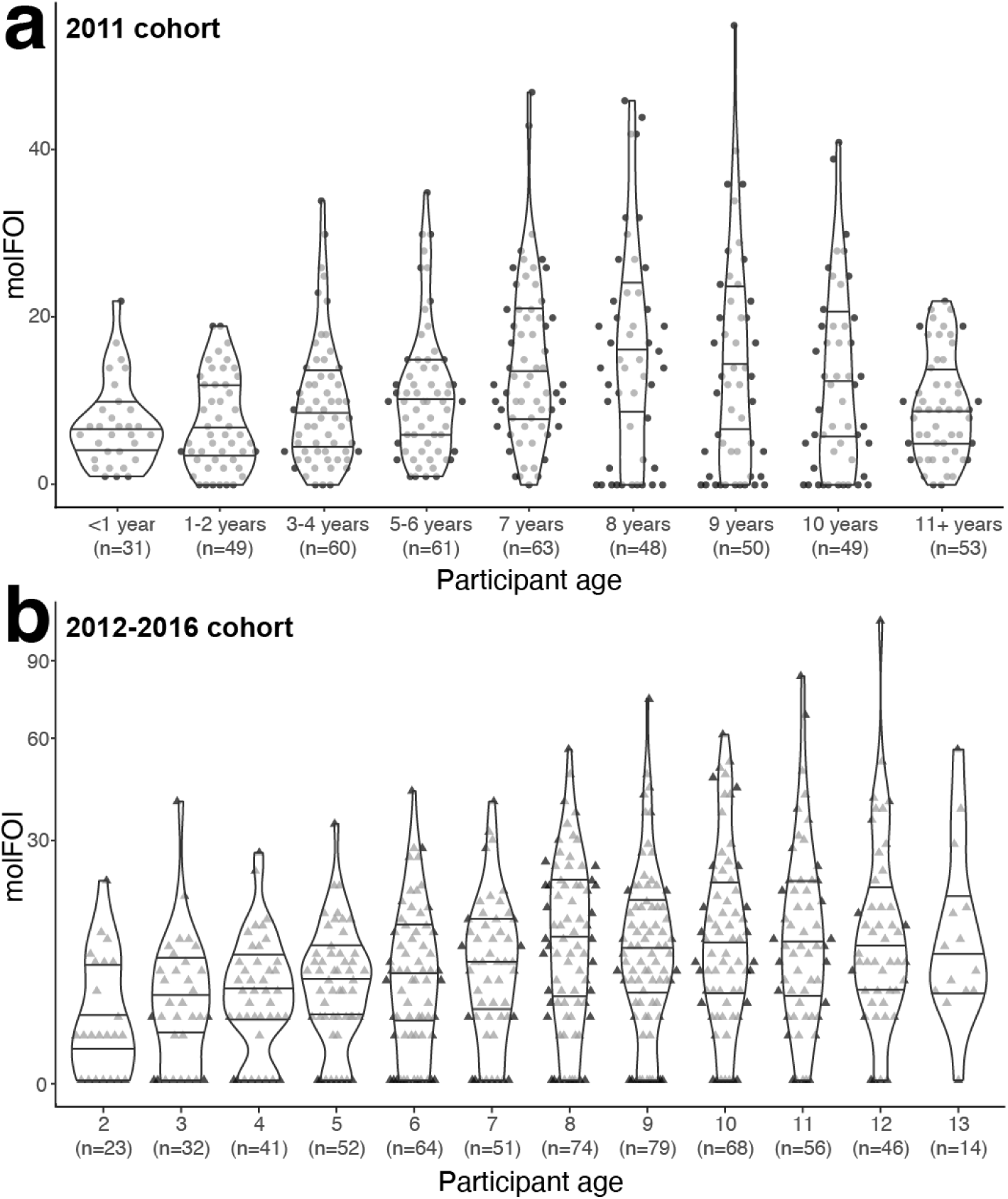
Molecular force of infection ranges in participants of all ages. (a) Each point represents a molFOI value; one for each of 464 participants over the 2011 season. The x-axis groups participants by age, aiming for similar sized age groups for comparisons. We found no significant association between age and molFOI (quasi-Poisson regression, p-value = 0.077). (b) Each triangle represents a molFOI value for one participant over a single transmission season, from 2012-2016. Each participant has five molFOI values in total, with increasing age. We did have a significant p-value for a linear regression (linear regression, adjusted R^2^ = 0.076, p-value = 4.4e^-12^ or quasi-Poisson regression, p-value = 4.8e^-13^), but a linear mixed-effects model with participant identity as a random effect fit the data better (likelihood rank test, □^2^(1) = 170.69, p-value < 2.2e^-16^). The pseudo-R^2^ value for all effects of 0.50, while the pseudo-R^2^ value for fixed effects only (age, in this model) was 0.00. In both plots,violin plots represent the density distributions, with horizontal lines at the 25th, 50th, and 75th percentiles. Abbreviations: molFOI, molecular force of infection.

We performed a similar analysis for the 2012-2016 dataset, except that we estimated molFOI for each participant per season, resulting in five molFOI values per participant (**Figure 2b**; participant ages reflect the increasing age per season.) These molFOI per season values ranged from 0 to 106 (mean: 10, median: 6, standard deviation: 12). We found a significant association between molFOI and participant age (linear regression, adjusted R^2^ = 0.076, p-value = 4.4e^-12^ or quasi-Poisson regression, p-value = 4.8e^-13^), but a linear mixed-effects model with age as a fixed effect and participant identity as a random effect fit the data better than the age-only model (likelihood rank test, □^2^(1) = 170.69, p-value < 2.2e^-16^). In the mixed effects model, fixed effects (participant age) had a pseudo-R^2^ value of 0.00, while the pseudo-R^2^ value for all effects was 0.50, suggesting that participant identity is a more significant driver of variation in infection risk than age.

### Intra-individual molFOI is consistent across seasons

We next compared individual molFOI across transmission seasons to determine whether molFOI variation in 2011 was driven by stochastic factors, or whether differences in infection risk among participants are consistent over time. We hypothesized that while heterogeneity in infection risk might obscure age-associated differences when comparing age bins cross-sectionally, these differences might be evident when profiling individuals longitudinally. We estimated the cumulative molFOI over the subsequent five seasons (2012-2016) and compared that value to the value from 2011 for individual participants (**Figure 3a**). We rejected the null hypothesis that variation in 2011 molFOI is stochastic; participants with high molFOI in 2011 generally had high molFOI in 2012-2016 as well (Spearman’s rank correlation, ϱ(111) = 0.53 (95% CI = [0.38, 0.66]), p < .001). We next stratified the 2012-2016 data by season, to test for confounding factors due to potential variation in weather or other temporal factors (**Figure 3b**). We found no significant differences in molFOI stratified by season (Kruskal-Wallis test, □^2^(4) = 4.36, p-value = 0.36). We visualized the patterns of average molFOI per age in each season (**Figure 3c**). We found that certain sub-cohorts, such as those aged 9 in 2012, consistently had high average molFOI values over time, despite their increasing age and cumulative parasite exposure. We also examined infection status in May 2011 (at study enrollment) as a predictor of infection risk; 44% of participants (n=202) were asymptomatically infected at enrollment. We found that molFOI from the 2011 season was significantly greater for participants who were infected at enrollment (**Figure 3d**, mean molFOI for infected = 15.9; uninfected = 7.6; Kruskal-Wallis test, □^2^(1) = 77.59, p-value < 2.2e^-16^). We also considered bednet usage as a potential explanation for molFOI variability, but participants universally reported daily bednet usage when surveyed in 2013 (561/563 households owned bednets and 559/563 households reported daily usage).

**Figure 3.**
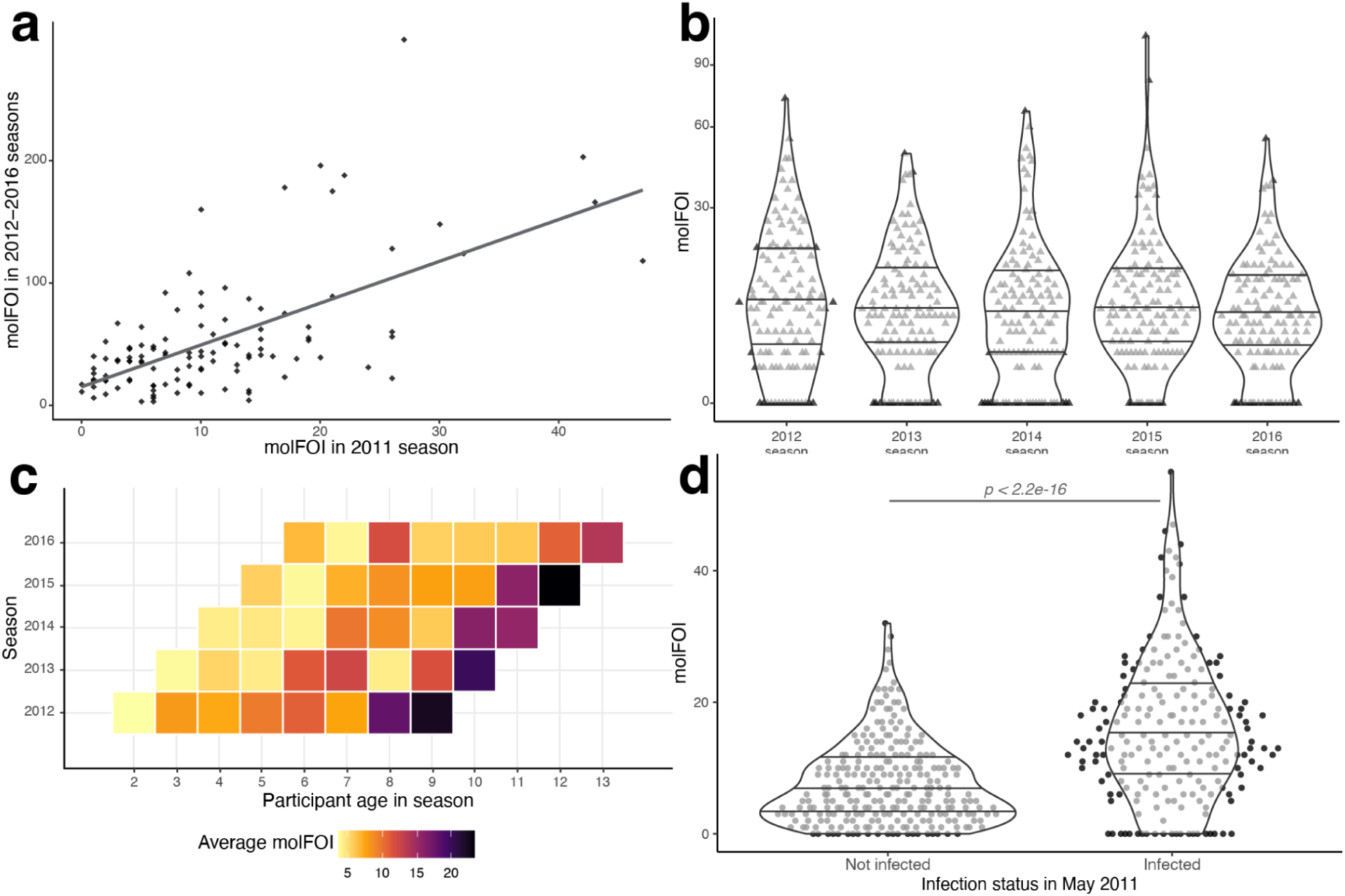
molFOI varies by participant, not by age. (a) Each point represents one of the 120 participants included in both of our datasets, showing their molFOI from the 2011 season (x-axis) and over the 2012-2016 seasons as a whole (y-axis). The axis scales vary due to the difference in time periods and sampling strategy between the two datasets (see Figure 1). The line represents a linear regression (adjusted R^2^ = 0.38, p-value = 3.4e^-13^). We also computed the Spearman’s rank correlation between molFOI in 2011 and 2012; we found a highly significant, positive correlation (⍴(111) = 0.53 (95% CI = [0.38, 0.66]), p < .001). (b) molFOI values for all participants for each of the five seasons included in the 2012-2016 dataset. The y-axis is on a squared scale for increased visibility of the lower molFOI values. No significant differences between groups (Kruskal-Wallis test, □^2^(4) = 4.36, p-value = 0.36). (c) Each tile is colored by the average molFOI of all participants of a given age (on the x-axis) during a given season (on the y-axis). (d) molFOI from the 2011 season, stratified by infection status at enrollment into the study, in May 2011. molFOI for participants who were infected at enrollment is significantly higher than for those not infected at enrollment (Kruskal-Wallis test, □^2^(1) = 77.59, p-value < 2.2e^-16^). Violin plots in (b) and (d) represent the density of the distributions, with horizontal lines at the 25th, 50th, and 75th percentiles. Abbreviations: molFOI, molecular force of infection.

We compared infection risk to rates of symptomatic disease by evaluating molFOI with respect to the number of treatments per participant within a transmission season or the parasitemia of individual samples (**Extended Data Figure 2**). We found that the number of treatments (a proxy for disease risk) had a significant association with molFOI (quasi-Poisson model, t(462) = 3.07, p = 0.0023), but this model explained only 2.02% of deviance in 2011 molFOI values, suggesting that disease risk does not capture much of the observed variation in infection risk. We also hypothesized that lower density infections would correlate with lower molFOI, if participants had parasitemia near the level of detection. However, participants grouped by low, middle, or high molFOI in 2011 had no differences in parasite densities (Kruskal-Wallis test, □^2^(2) = 4.68, p-value = 0.096). In fact, participants with high molFOI in 2012-2016 had slightly lower densities (mean densities: 2.5e^4^ (low molFOI), 2.1e^4^ (mid molFOI), 2.4e^4^ (high molFOI); Kruskal-Wallis test, □^2^(2) = 10.52, p-value = 0.0052; Dunn test with Holm correction, p-value = 0.0037 (low vs. high)), rejecting our null hypothesis. Individuals with more anti-parasite immunity could have lower parasite densities, and perhaps they are more likely to accumulate multiple clones over the course of lower density, potentially asymptomatic infections.

### Host factors explain some heterogeneity in molFOI

We next hypothesized that host-specific factors could affect infection risk. We found that participant sex was not associated with molFOI in 2011 (**Figure 4a**, mean molFOI for females = 11.6, males = 11.1; Kruskal-Wallis test, □^2^(1) = 2.47, p-value = 0.12) or 2012-2016 (**Figure 4b**, mean cumulative molFOI for females = 47.7, males = 54.0; Kruskal-Wallis test, □^2^(1) = 0.77, p-value = 0.38). Knowing that many individuals in this population carry malaria-protective variant alleles at the hemoglobin subunit beta locus (HBB)^11^, we examined molFOI stratified by HBB genotype (**Figure 4c,d**). Of the 464 participants in our 2011 cohort, 372 were homozygous for the ancestral allele, A (HbAA genotype). A total of 47 were heterozygous for the C allele (HbAC genotype), which has been associated with lower risk of severe malaria disease^13,14^. A total of 43 participants were heterozygous for the S allele (HbAS genotype), which confers the sickle cell trait and has been associated with lower risk of both uncomplicated and severe malaria disease^13,15,16^. (We excluded 2 participants with HbCC and HbSC genotypes from the analysis in **Figure 4c** only). We found significant differences in the molFOI in 2011 for participants with HbAS compared to both HbAA and HbAC genotypes (**Figure 4c**, mean molFOI for HbAA (11.8), for HbAC (13.1), for HbAS (5.3). Kruskal-Wallis test, □^2^(2) = 23.53, p-value = 7.8e^-6^, ε^2^ = 0.05. Dunn test with Holm adjustment, p-values = 7.6e^-6^ (HbAA vs. HbAS), 9.1e^-5^ (HbAC vs. HbAS), 0.51 (HbAA vs. HbAC)). Of the 120 participants in our 2012-2016 cohorts, fewer had variant genotypes (6 HbAC and 17 HbAS); molFOI values were lower in HbAC (mean molFOI: 45.5) and HbAS (mean molFOI: 34.3) participants than in HbAA participants (mean molFOI: 54.3), but they did not reach the threshold of significant difference (**Figure 4d**; Kruskal-Wallis test, □^2^(2) = 0.96, p-value = 0.62).

**Figure 4.**
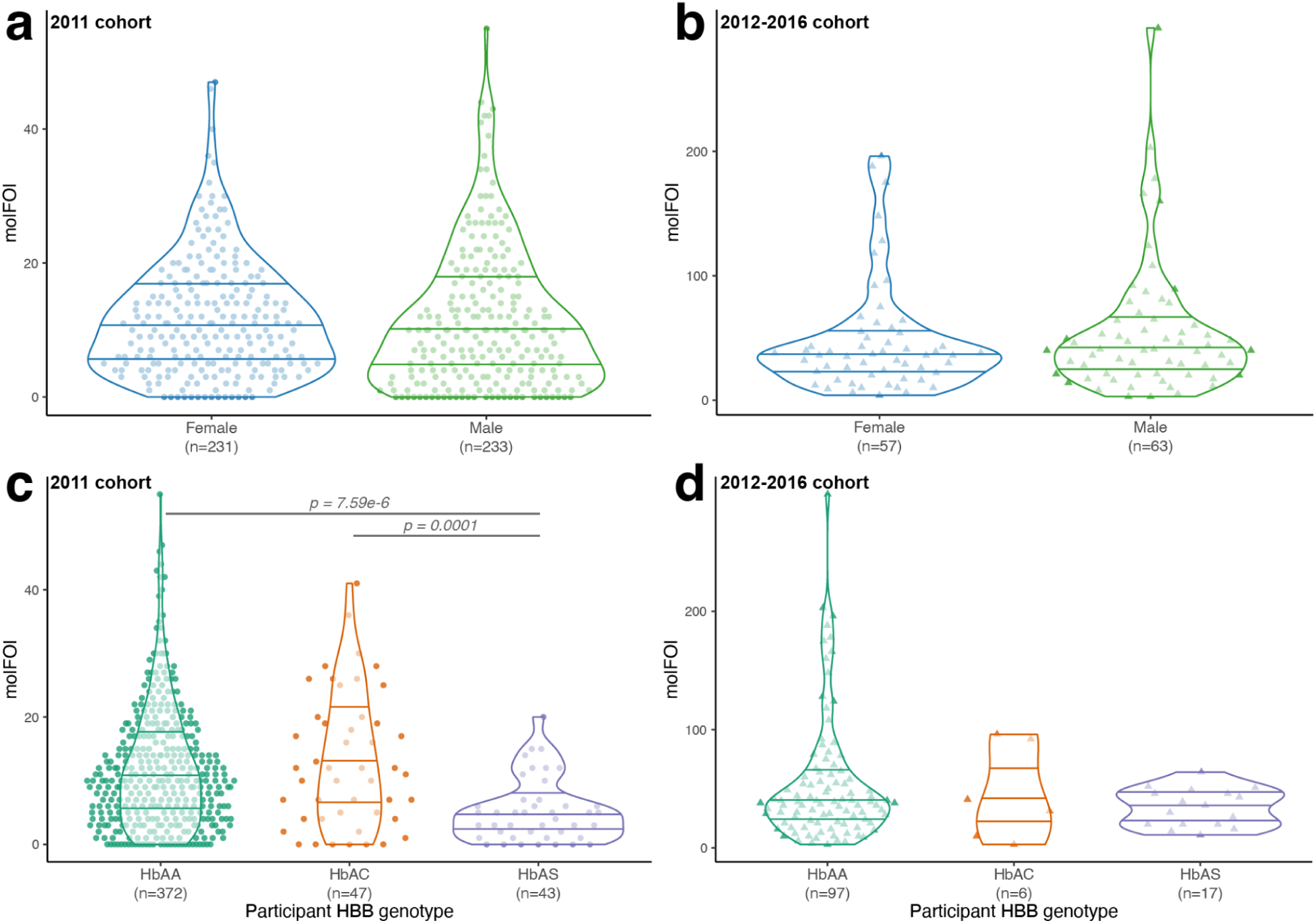
Host genotype explains some variability in molFOI, while participant sex does not. (a) molFOI values from 2011, stratified by participant sex. No significant differences between groups (Kruskal-Wallis test, (a) □^2^(1) = 2.47, p-value = 0.12). (b) molFOI values from 2012-2106, stratified by participant sex. No significant differences between groups (Kruskal-Wallis test, □^2^(1) = 0.77, p-value = 0.38). (c) molFOI values from 2011, stratified by participant HBB genotype. Horizontal lines mark significant differences between groups (Kruskal-Wallis test, □^2^(2) = 23.53, p-value = 7.8e^-6^, ε^2^ = 0.05. Dunn test with Holm adjustment, p-values = 7.6e^-6^ (HbAA vs. HbAS), 9.1e^-5^ (HbAC vs. HbAS), 0.51 (HbAA vs. HbAC)). (d).molFOI values from 2012-2016, stratified by participant HBB genotype. No significant differences between groups (Kruskal-Wallis test, □^2^(2) = 0.96, p-value = 0.62). In all panels, colors represent the same categories as the x-axis. Violin plots represent the density of the distribution, with horizontal lines representing the 25th, 50th, and 75th percentiles. Abbreviations: HBB, hemoglobin subunit beta locus; molFOI, molecular force of infection.

### Geographic features may explain some heterogeneity in molFOI

We next considered whether geographic factors could affect infection risk. First, we analyzed the spatial autocorrelation of the 2011 molFOI values (**Figure 5a**), to look for potential hotspots, or geographic regions with patterns of molFOI. We found a very small dispersion of molFOI (Moran’s I = -0.023, pseudo-p-value = 0.002, two-sided hypothesis, compared to a Monte-Carlo simulation of permutations of the data). We found a slight positive correlation between the distance from participants’ homes to the Kalifabougou study clinic and their molFOI in 2011 (**Figure 5b**, quasi-Poisson regression, t(312) = 3.00, p = 0.003). We also found a small positive correlation between participants’ distance to the closest part of the river system and their 2011 molFOI (**Figure 5c**, quasi-Poisson regression, t(312) = 5.71, p = 2.68e^-8^). Taken together, these spatial analyses suggest that geographic differences may have a slight effect, but they do not explain the majority of the observed heterogeneity in infection risk in this cohort (2.45% and 8.30% of deviance explained for distance to clinic and rivers, respectively).

**Figure 5.**
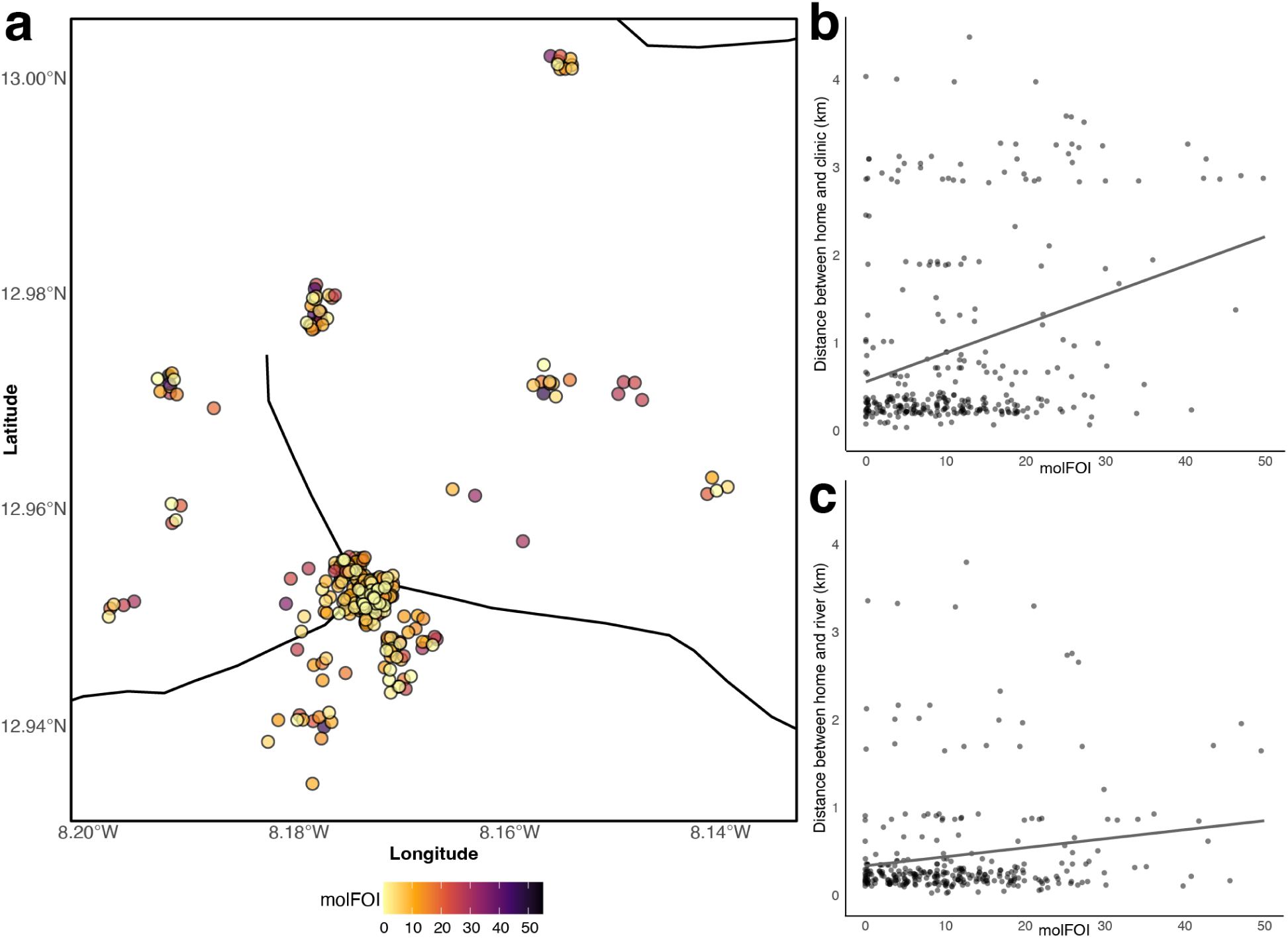
molFOI does not concentrate in geographic hotspots. (a) Visualization of homes and molFOI in Kalifabougou, Mali. Each dot represents the geographic coordinates of one participant’s home, colored by their molFOI in 2011. Points are slightly jittered, for visibility. Black lines represent local rivers, geospatial data from Global Map of Mali © ISCGM/IGM. (b) Scatter plot of each participant’s molFOI in 2011 (x-axis) vs. the distance from their home to the Kalifabougou clinic (y-axis, labeled in kilometers) (quasi-Poisson regression, t(312) = 3.00, p = 0.003). The line shows the linear regression (adjusted R^2^ = 0.089, p-value = 4.3e^-8^). (c) A similar scatter plot to panel b, except that the y-axis shows the distance from each participant’s home to the nearest river (quasi-Poisson regression, t(312) = 5.71, p = 2.68e^-8^). Linear regression shown as described above (adjusted R^2^ = 0.025, p-value = 0.0031). Abbreviations: km, kilometers; molFOI, molecular force of infection.

### Several serological features distinguish between low and high infection risk participants

Systems serological profiling was performed on 201 participants overlapping with our sequencing dataset^12^. These data were generated using samples from May 2011, at enrollment into the study and before the transmission season began that year, with the objective of defining potential correlates of protection. Antigen-specific IgG1, IgG2, IgG3, IgG4, IgA1, and IgM to AMA1, CSP, MSP1, RH5 and the N-and C-terminal domains of CSP were quantified. The functional potential of these antigen-specific antibodies was also captured, including the ability of the antigen-specific antibodies to bind to Fc receptors (FcRn, FcγRIIAH, FcγRIIAR, FcγRIIB, FcγRIIIAF, FcγRIIIAV, and FcγRIIIB) and recruit antibody-dependent complement deposition, antibody-dependent cellular phagocytosis, and antibody-dependent neutrophil phagocytosis. While the evolution of antigen-specific neutrophil activation correlated with rates of disease severity^12^, here we aimed to define whether malaria-specific serological markers were positively or negatively associated with molFOI.

We set thresholds for molFOI in 2011 to create “low molFOI” (<4) and “high molFOI” (>13) groups (**Figure 6a**). To explore the sensitivity of the analysis to threshold used, we defined three different sets of thresholds, to consider the upper and lower 33%, 25%, and 12% of the molFOI distribution (**Extended Data Table 1**). We primarily discuss results from the middle set of thresholds here; the others are highly concordant and are described in **Extended Data Figure 3** and **Extended Data Figure 4**.

**Figure 6.**
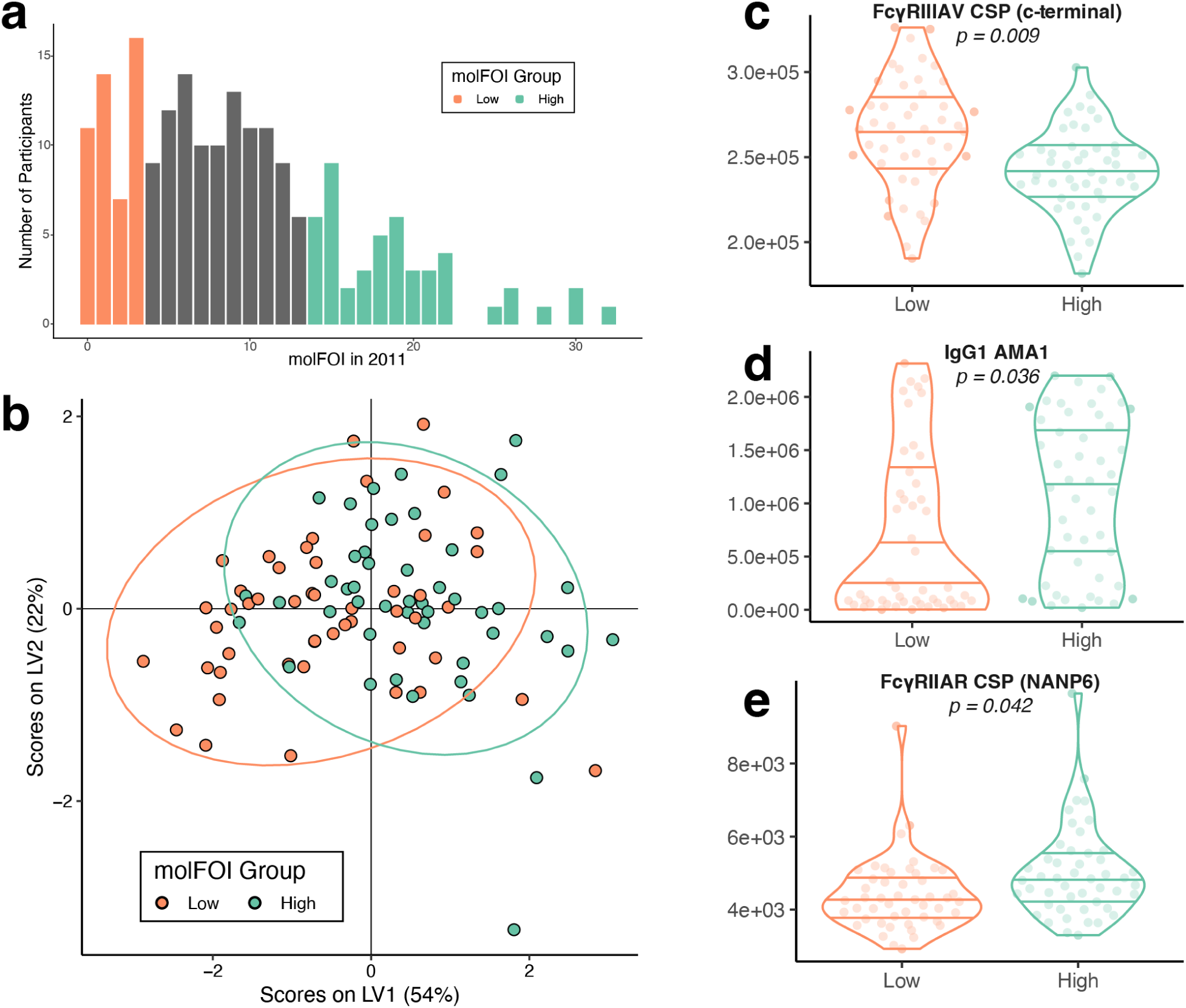
Immunological features distinguish between high and low molFOI participants. (a) Distribution of molFOI in the 2011 season for the 201 participants for whom we had both genetic and serology data. We compared low and high molFOI participants in the following analyses, who are colored in orange and teal, respectively. (See **Extended Data Table 1** for details on molFOI ranges used here and in other versions of this analysis). (b) We used the LASSO algorithm to select features that stratified the low and high molFOI groups. This plot shows the partial least squares latent variable scores based on the selected features. (c-e) Raw data from the 3 selected features that were selected by the LASSO algorithm. Each point represents a single participant. p-values are from Mann-Whitney tests, with Benjamini-Hochberg corrections. Violin plots represent the density of the distribution, with horizontal lines representing the 25th, 50th, and 75th percentiles. Abbreviations: AMA1, apical membrane antigen; IgG, immunoglobulin G; LV, latent variable; molFOI, molecular force of infection; NANP6, repeat region within circumsporozoite protein.

We used the least absolute shrinkage and selection operator (LASSO) algorithm to select immune features that differentiated the two groups, and we used partial least squares discriminant analysis (PLS-DA) to build classifiers. These classifiers distinguished between groups (**Figure 6b**) with an accuracy of 66%. We performed ten-fold cross-validation of all classifiers, with two different null models: one with permuted group labels and one with random feature selection in place of LASSO (**Extended Data Figure 5**). The 66% accuracy of the PLS-DA classifier was above the 95th percentile of the accuracy of both types of cross-validation (**Extended Data Figure 5cd**). We also looked at the features that were selected via LASSO and included in the classifier (**Figure 6cde**). Fc receptor binding to the highly polymorphic c-terminal region of CSP (specifically, Fcγ receptor IIIAV, or FcγRIIIAV) was enriched in participants with low molFOI (Mann-Whitney test with Benjamini-Hochberg correction, p-value = 0.009). Interestingly, Fcγ receptor IIAR binding (FcγRIIAR), to a distinct part of CSP, the NANP repeat region, was enriched in participants with high molFOI (Mann-Whitney test with Benjamini-Hochberg correction, p-value = 0.042), along with AMA1-specific IgG1 responses (Mann-Whitney test with Benjamini-Hochberg correction, p-value = 0.036). We also compared the features that were selected in multiple instances of the threshold specifications that we performed (**Extended Data Table 1**). We found that FcγRIIIAV binding to the c-terminal of CSP was enriched in participants with low molFOI in all three specifications of thresholds, suggesting that it may be protective against infection. IgG1 specific to AMA1 was enriched in high molFOI participants in two of the three threshold specifications, similar to previous studies suggesting that it may be a marker of infection history^17–19^.

## Discussion

In this study, we identified an unexpectedly large degree of malaria infection risk heterogeneity among individuals in a small community. This heterogeneity in infection risk was consistent over time, and was only partially explained by measured geographic, demographic, behavioral, and host genetic factors. Heterogeneity in infection risk has likely been previously under characterized in most studies, and was measured with unprecedented precision in this study through multiplexed amplicon genotyping of a very densely sampled longitudinal sample collection from 464 participants.

Symptomatic malaria incidence is only weakly associated with molFOI in this cohort, suggesting that some previous symptomatic disease-based studies^20,21^ have not captured the true magnitude of infection risk heterogeneity in endemic settings. We found that molFOI exhibits a large degree of variation among participants (2011 range: 0-55, standard deviation: 9), and remains consistent within participants over time. Participants with high molFOI in 2011 were likely to have high molFOI in subsequent seasons as well, regardless of their increasing age and presumed development of natural immunity. This finding suggests that the variation in infection risk that we observed within individual transmission seasons is not stochastic, not largely attributable to NAI, and is driven by individual features (**Figure 3**). We found no significant differences in infection risk by participant sex (**Figure 4ab**), although recent work in a different cohort has found increased risk within school-age males^22^. We did find that participants with asymptomatic infections at study enrollment had higher molFOI, consistent with a recent genotypic analysis of the RTS,S/AS01_E_ fractional dosage trials, in which participants who were infected at baseline had higher molFOI in the first 2 months of the study^23^.

Our data indicate that host genotype may be one modulator of infection risk. Participants from this cohort were previously genotyped at the hemoglobin subunit beta (HBB) locus, given the known protective effects of the sickle (HbS) and HbC alleles against severe disease^13^ Similar to a previous finding, which estimated force of infection using capillary electrophoresis genotyping^24^, our finding of lower molFOI in HbAS participants than HbAA or HbAC (**Figure 4c**) may indicate that HbAS participants are protected against blood-stage infection, and/or that blood stage infection duration was shorter and less likely to be detected. However, HbAS participants only represent 10% of the cohort participants, and the effect size of this association was small (ε^2^ = 0.05). Despite HBB’s role as the most important locus conferring protection against clinical malaria^13^, it only explains a small portion of variation in infection risk in this cohort. Other unmeasured host genetic factors may also influence infection risk, but are likely to exhibit smaller effect sizes than HbS and HbC.

Previous studies have linked infection risk heterogeneity with geographic factors^25,26^. We found significant but small associations between molFOI heterogeneity and distance to the central clinic in Kalifabougou, as well as distance to the nearest river (**Figure 5**), suggesting that geographic factors may contribute a small amount to the heterogeneity of infection risk we observed. Studies with a wider range in geographic and ecological factors^27^ than this one may benefit from considering geographic factors. Behavioral factors, including bednet usage, have also been indicated in previous studies of disease risk heterogeneity; however, participants in this cohort almost universally reported daily bednet usage.

We conducted a systems serology analysis to identify candidate immune correlates of infection risk vs. infection protection, using molFOI as a composite measure of disease risk/protection. Analysis of systems serology data from a subset of participants profiled in this study identifies several potential correlates of infection risk or protective immunity^12^. We found that participants with low molFOI in 2011 (low risk, and/or high protection) had enriched Fcγ receptor IIIA binding antibody responses to the C-terminus of CSP (**Figure 6c**). As CSP is expressed in sporozoites, an immune response to CSP could potentially prevent progression to blood-stage infection and reduce molFOI. This finding concords with recent studies of serological responses to the CSP-based RTS,S/AS01 vaccine that suggested a protective role for Fc-effector functions, particularly those specific to CSP^28,29^, as well as previous work with this cohort that found an association between functional CSP-specific IgG and protection from clinical disease and decreased parasite density^12^. A genotypic analysis of the phase 3 trial of RTS,S/AS01 found enhanced protective efficacy against infections exactly matching the vaccine strain in the CSP C-terminus, underscoring the role of this region in vaccine-induced immunity^30^. The importance of the CSP C-terminus for natural immunity is strongly indicated by extremely high levels of natural polymorphism, presumably generated by immune-mediated balancing selection^31^. Further work examining responses to different vaccine dose regimens found an increased response to CSP c-terminal-specific responses in participants who received the dose regimen that had higher vaccine efficacy^32^. Finally, a recent study examining responses to the radiation-attenuated whole sporozoite PfSPZ vaccine found increased expression of genes related to Fcγ receptor-mediated phagocytosis correlated with protective outcomes, regardless of vaccination status^33^. Taken together, these results suggest that increased response to CSP with antibodies able to bind FcγRIIIA and activate peripheral or liver-resident NK cells, phagocytic cells, and memory CD8+ T cells may be an important mechanism of immune protection for both natural and vaccine-induced immunity to infection, even if the implicated regions of CSP differ (C-terminus and NANP central repeat^29^, respectively)..

The systems serology data also identified several candidate correlates of infection risk rather than protection. We observed enriched responses to AMA1-specific IgG1 among participants with higher molFOI in 2011 (**Figure 6d**). Other studies have reported similar results, with positive associations between levels of antibodies to merozoite antigens (like AMA1) and risk of disease^19,34^ or infection^17,18^. In particular, some studies^10,17^ supported a previously proposed threshold model^35^, in which antibody levels may serve as markers of infection risk or protection, when they are below or above a protective threshold, respectively. In addition, we observed a positive association between IgG1 recognition of the NANP6 peptide of CSP and higher molFOI in 2011 (**Figure 6e**). Though IgG response to the NANP6 peptide is a strong correlate of protection for the RTS,S/AS01 vaccine^29^, this relationship may not hold for NAI.

This work has some limitations. Amplicon sequencing data recovery was variable across samples, with dropout of one or more amplicons in 35% (n=1,540) of 4,383 parasite-positive samples. We mitigated this by requiring conservative approaches to define new infections. The age of some of the dried blood spots from which we extracted DNA may have impacted our sensitivity to detect new infections in a systematic manner, but we observed consistent distributions of molFOI from samples from 2012-2016 (**Figure 3b**), despite a five-year difference in original sample age. Additionally, heterogeneity in the number of treatments a participant received over a period of time and the density of parasites present within samples have the potential to bias these analyses, by creating variable periods of time during which participants were refractory to new infections. We did find expected correlations between both parasite density and number of treatments with participant age (**Extended Data Figure 2**); older participants tended to have fewer treatments and lower density infections. However, if these factors were to bias our analyses, we expect that molFOI would decrease in participants with lower density infections and/or fewer treatments. Instead, we found slightly positive correlations -participants who were treated more often tended to have higher molFOI than those treated less. We found no difference in parasite density data for high vs low molFOI participants in 2011, though we did find a slightly lower density for participants with high molFOI in 2012-2016 than those with low molFOI. Overall, these differences trend in the opposite direction than what we hypothesized could bias our analysis, though the differences are small.

In summary, the significant inter-individual variation in infection risk we observed in this cohort study is largely not accounted for by diverse measured variables, motivating future studies to identify these factors. In particular, factors associated with risk of symptomatic disease in previous studies could play a role, particularly variability in mosquito biting rates among people^22,36–39^, or intrinsic immune factors like HLA genotype that may not be dependent on age and previous exposure^40^. Heterogeneity in infection risk should be considered in future clinical trials of malaria interventions, particularly in smaller clinical trials where randomization may not account for the wide range in risk and could confound interpretation of intervention efficacy. Additionally, interventions could have differing efficacy according to individual infection risk and associated immune response, as has been hypothesized or observed for several malaria vaccine trials^23,33,41,42^. Finally, an enhanced understanding of the drivers of infection risk heterogeneity could inform measures to alleviate risk and assist in targeting interventions to better protect those most vulnerable to infection, especially when deployment of interventions is constrained due to manufacturing limitations^43^ or shortfalls in local public health resources.

## Methods

### Samples from cohort

We processed samples from the Kalifabougou longitudinal cohort study previously described^7^ (**Figure 1**). The Kalifabougou cohort study was approved by the Ethics Committee of the Faculty of Medicine, Pharmacy and Dentistry at the University of Sciences, Technique and Technology of Bamako, and the Institutional Review Board of the National Institute of Allergy and Infectious Diseases, National Institutes of Health (NIH IRB protocol number: 11IN126; https://clinicaltrials.gov/; trial number NCT01322581). The original study authors obtained written informed consent from parents or guardians of participating children before inclusion in the study, and they collected these samples between 2011 and 2016. We sequenced samples from 464 participants of the study’s full 695 participants who were included in the 2011 cohort, prioritizing a range of ages and even distribution of participant sex. We also selected samples from 120 participants who were enrolled in further study from 2012-2016, prioritizing younger ages, even participant sex distribution, and number of samples available per participant.

### DNA extraction and sequencing

We physically randomized all samples prior to DNA extraction, to minimize the impact of any batch effects. We extracted DNA from the clinical samples and sequenced them using the 4CAST amplicon panel, as previously described^44^. We sequenced samples from 2011 on Illumina MiSeq instruments, with 96-384 samples within each run. We sequenced samples from 2012-2016 on Illumina NovaSeq instruments, with 768 samples per lane. Data from these samples were submitted to the NCBI Sequence Read Archive (http://www.ncbi.nlm.nih.gov/sra) under accession PRJNA1129562.

### Data processing

We processed the paired-end sequencing data through a custom pipeline^44^, based on the Divisive Amplicon Denoising Algorithm (DADA2)^45^, which produces pseudo-CIGAR strings summarizing the observed polymorphisms in each haplotype. In the 2011 dataset, where the number of samples varied per sequencing batch, we divided the read-pairs at this stage by 2 or 4, for batches of 192 or 96 samples, respectively. Next, we required a haplotype to have a minimum of 10 reads per sample, as well as a minimum of 1% of the within-sample reads per locus. Finally, we removed any haplotypes that only appeared once in the entire dataset. In the 2012-2016 dataset, which we sequenced at higher depth, we required a haplotype to have a minimum of 200 reads per sample, with the same within-sample frequency and singleton haplotype removal as before.

### Defining molecular force of infection

We defined individual clones (individual haplotypes at any locus) and infection events (all haplotypes within a sample). To allow for imperfect data, stochastic dropout of loci, and sequestration of parasites, we tested different conditions to allow for clones to disappear for short periods of time and still be considered part of an ongoing infection. In the 2011 dataset, we tested the number of “skipped visits,” the number samples without a given clone before it reappears (0 - 4), the number of days between appearances of a given clone (15, 30, 60), and combinations of the two metrics. Although the number of new clones changed with this sensitivity testing, the overall conclusions of our analyses did not; thus, we defined clones as new if they are not detected for more than two visits in a row or more than 30 days. We also tested the sensitivity of our definitions of new infection events, requiring 1, 2, or 3 new clones present within a sample, or requiring new clones at 2 different loci present within a sample. We defined new infection events by the presence of 2 new clones within a sample. We also repeated a subset of the above combinations with a higher minimum read-threshold (50 read-pairs, instead of 10). Finally, we repeated all sensitivity tests with a higher threshold to retain haplotypes in the analyses: 50 read-pairs per haplotype per sample. We retained the 10 read-pair minimum threshold, and all analyses include data from all four loci, unless otherwise stated.

We repeated these sensitivity analyses on the 2012-2016 dataset, with only a few changes. Due to the change in sampling frequency in these years (biweekly to monthly), we increased the number of days between clone appearances in our tests (30, 60, or 90 days), and we defined clones as new if they are not detected for more than two visits in a row or more than 60 days. We also tested a higher read threshold here (500 read-pairs per haplotype per sample) and retained the 200 read-pair threshold.

We defined the molecular force of infection as the total number of new clones present within a participant over a given period of time (generally one transmission season). To do this, we looked at all new infection events for a participant and filtered to only new clones present then, as opposed to clones carried over from the previous visit. We then counted the maximum number of new clones present at any one of our four sequenced loci. Finally, we summed these maximum numbers of new clones present from each new infection event over the time period of interest.

### Spatial analysis

We used latitude and longitude coordinates of participant households, as well as the metrics of distance to clinic or water from these households, from earlier studies of this cohort^11^. We used the latitude and longitude coordinates for visualization purposes (**Figure 4a**), along with coordinates of local water (Global Map of Mali © ISCGM/IGM). We performed the analyses in **Figure 5bc** using the distance data from the previous cohort study. We used the R packages “sf” and “tidyverse” for the visualization in **Figure 5a**^46–48^. We used the “mat2listw” and “moran.mc” functions from the “spdep” R package for the Moran’s I analysis^47^.

### Systems serology analysis

These analyses included the 201 participants that overlapped between the serological data and the genetic data. We centered and scaled all data (Z-scored), using the “scale” function in R. We then created 3 different thresholds for selecting high and low molFOI participants (see **Extended Data Table 1**). We performed the same analyses on each subset of data. We limited our analysis to features that were significantly different between the two groups on their own, using Mann-Whitney tests with a Benjamini-Hochberg correction for multiple hypothesis testing. We then used the “ropls” and “systemsseRology” packages in R^49,50^; specifically, we used the “select_lasso” function to choose features significantly associated with molFOI via the LASSO feature selection algorithm. We ran this selection 100 times and retained features that were selected in at least 80 of the trials for building PLS-DA classifiers. We used the “train_ropls”, “predict_ropls”, and “score_accuracy” functions to assess the classifiers’ performance. We also used the “validate_repeat” function in a 5-fold cross-validation. We compared this performance to two different negative controls - one in which we permuted the “low molFOI” and “high molFOI” group labels before running the LASSO feature selection, and a second in which we kept the original group labels but selected random features instead of performing feature selection. We performed ten rounds of cross-validation, which included ten permuted group label trials and ten random feature selection trials per round.^12^

### Statistical analysis

We used R 4.1.2^51^ for all analyses, unless otherwise stated, with the packages “tidyverse,” “here,” and “RColorBrewer”^48,52,53^. We tested for significance using “glm” or “kruskal.test,” as described in figure legends. We used the “dunn.test” R package to perform Dunn tests with Holm post-hoc corrections for all analyses with multiple hypothesis testing, unless otherwise stated^54^. We used “cor_test” and “cor_to_ci” from the “correlation” package for the Spearman’s ranked correlation analysis in **Figure 3A**^55^. We used the “lmer” function from the “lme4” R package^56^ to fit linear mixed-effects models; we then used the “anova” function and the pseudo-R^2^ procedure as previously described^57^ to compare models. We used “geom_quasirandom” from “ggbeeswarm” for all jittered beeswarm plots^58^. We used Adobe Illustrator 2024 to create **Figure 1** and **>Extended Data Figure 1**, to arrange panels in all figures, and add lines to denote significant comparisons between groups (**Figure 3d**, **Figure 4c**, **Extended Data Figure 2e**).

## Data Availability

All sequencing data will be available through the NCBI Sequence Read Archive (http://www.ncbi.nlm.nih.gov/sra) under accession PRJNA1129562. The code used to process sequencing data is available at https://github.com/broadinstitute/malaria-amplicon-pipeline.

## Acknowledgements

We thank the participants in the study and their caregivers for making this study possible. We thank the Kalifabougou clinic and study team. We thank Charlotte Switzer for helpful discussions. We thank Shanping Li for help with the original cohort samples. This project was supported by an NIH R01 award to DEN (R01AI141544). The original Mali cohort study was funded by the Division of Intramural Research, National Institute of Allergy and Infectious Diseases, National Institutes of Health.

## Data availability

All sequencing data were submitted to the NCBI Sequence Read Archive (http://www.ncbi.nlm.nih.gov/sra) under accession PRJNA1129562.

## Code availability

The code used to process sequencing data is available at https://github.com/broadinstitute/malaria-amplicon-pipeline.

## Author contributions

● Conceptualization EL, COB, PDC, TMT, DEN
● Formal Analysis EL
● Funding Acquisition COB, DEN
● Investigation ZMJ, MS, NN
● Methodology EL, ZMJ, MS
● Project Administration DEN
● Resources NN, GA, PDC, BT, TMT
● Supervision COB, GA, DEN
● Validation EL, ZMJ, MS
● Visualization EL
● Writing - Original Draft Preparation EL, DEN
● Writing - Review & Editing [All authors]

## Extended Data

**Extended Data Table 1.**
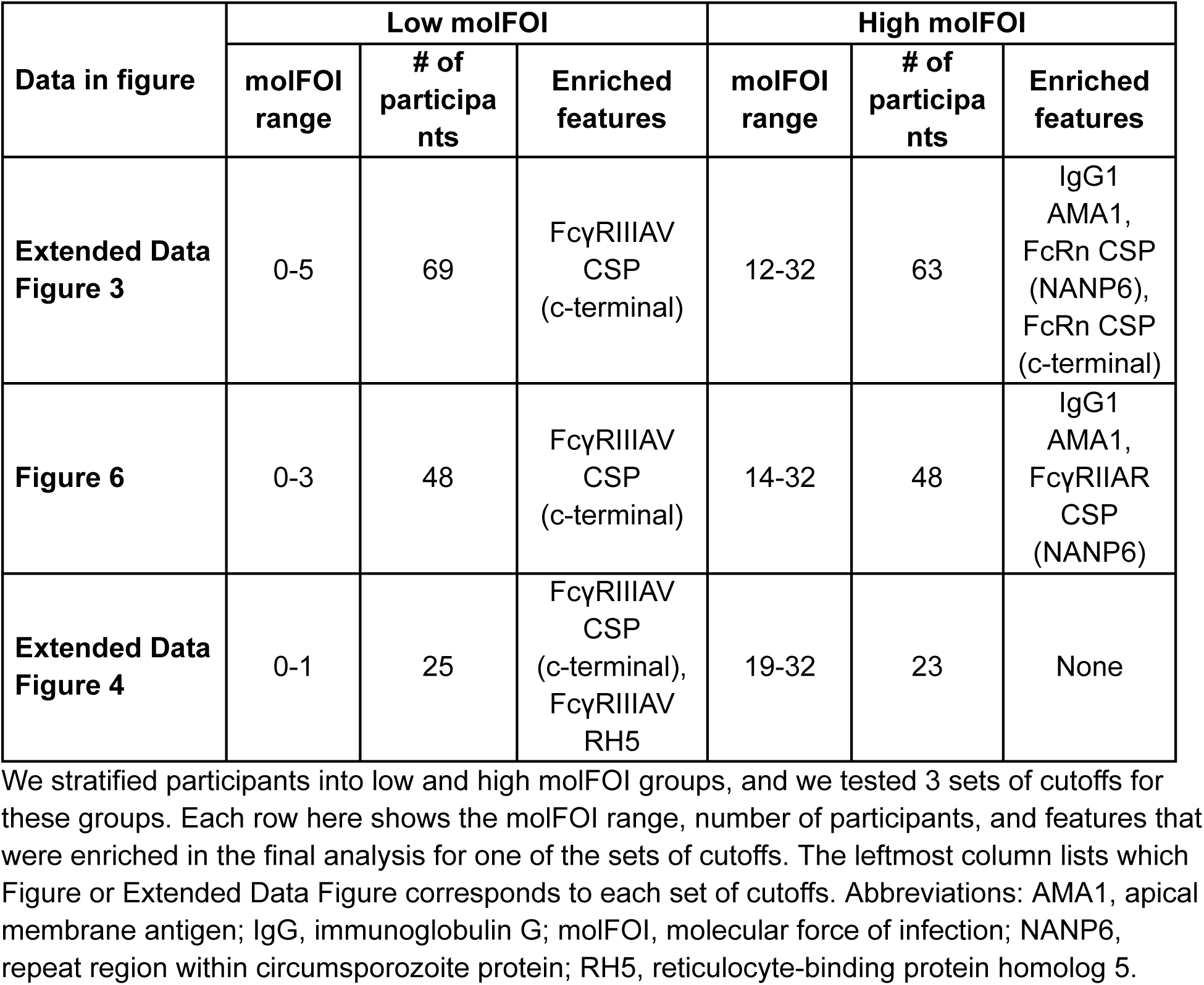
Immunological features enriched in participants with high and low molFOI.

**Extended Data Figure 1.**
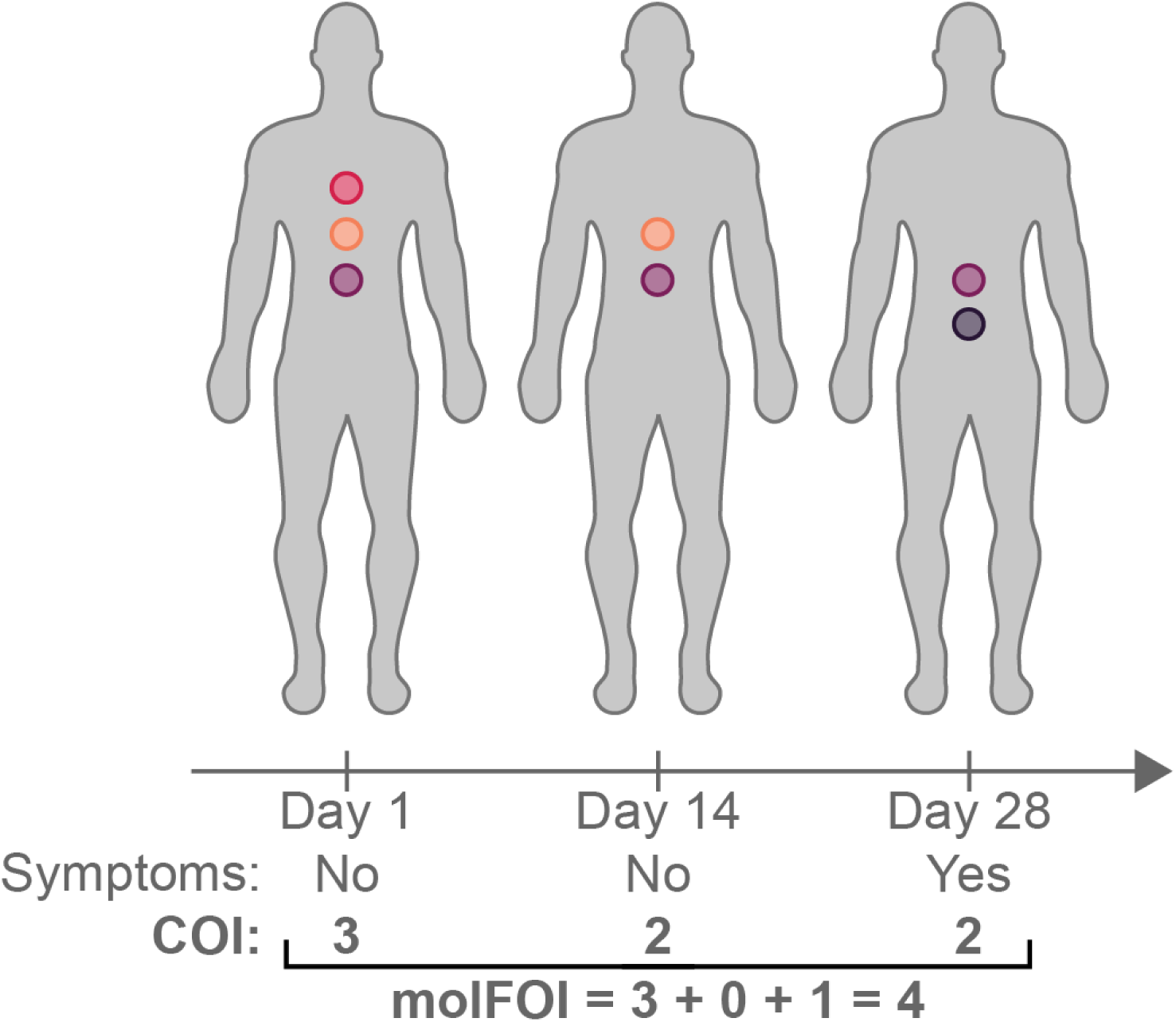
Molecular force of infection and other metrics of parasite dynamics. This example shows the parasite dynamics within a single person over a 28-day period. Each circle within a person above represents a detectable parasite clone. The pink and orange parasites are both from asymptomatic infections; the pink parasite only appears once, while the orange parasite is an ongoing asymptomatic infection. The purple and black parasites represent symptomatic infections. The purple parasite represents an infection that transitions from asymptomatic to symptomatic, and the black parasite is immediately symptomatic. Regardless of infection type of the individual clones, we estimate the complexity of infection (COI) at each time. We also summarize the number of new infections over a period of time by using molFOI, the molecular force of infection. This counts the new COI at each infection, while not including clones that continue from an already-accounted-for infection. Abbreviations: COI, complexity of infection; molFOI, molecular force of infection.

**Extended Data Figure 2.**
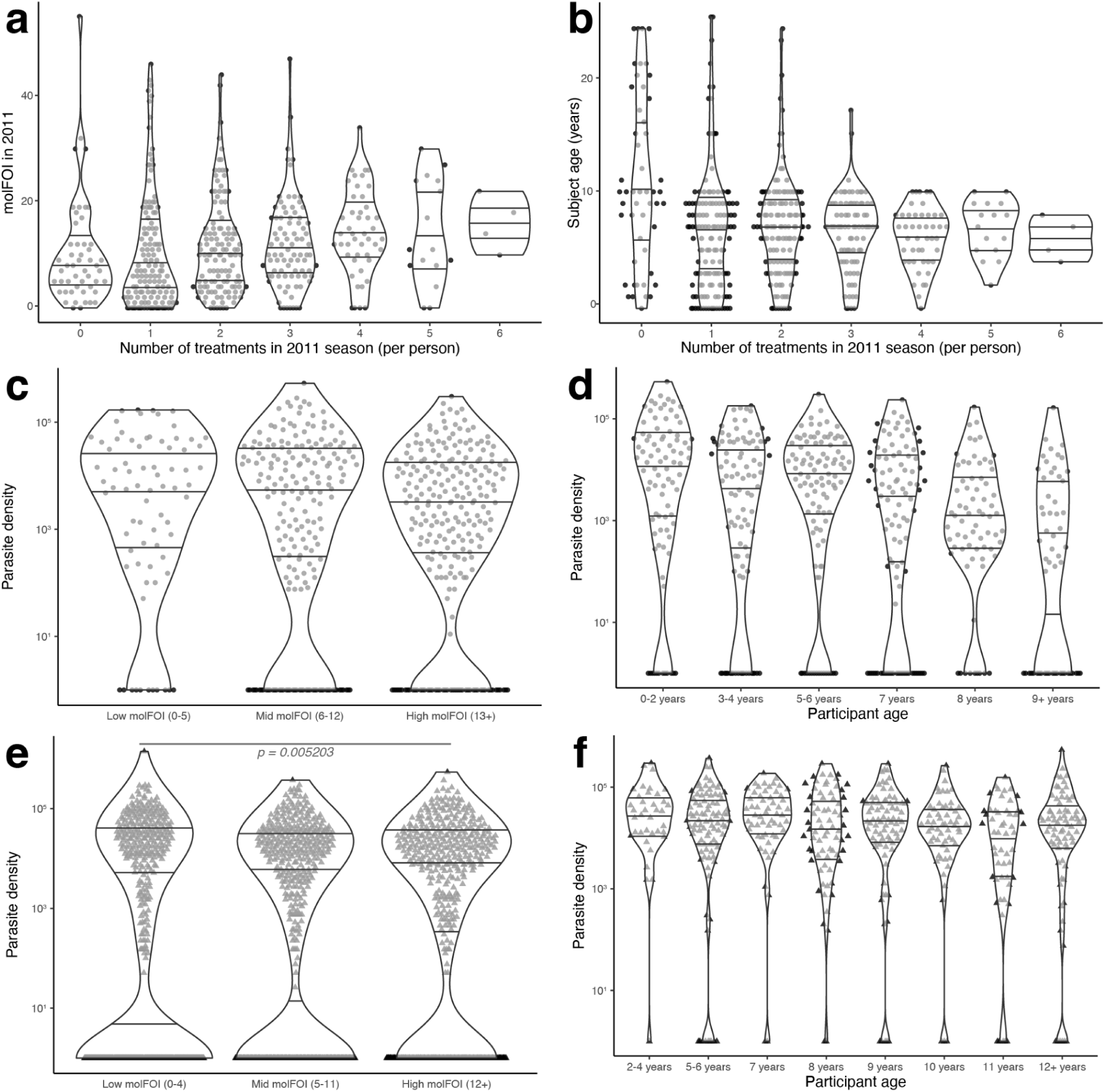
Heterogeneity of number of treatments and parasite density. We compared the number of treatments per person in the 2011 season with the participants’ molFOI over the 2011 season (a) and the participants’ ages at enrollment into the study (b). We found a significant interaction between number of treatments and 2011 molFOI (quasi-Poisson model, t(462) = 3.07, p = 0.0023, 2.02% of variance explained), and we found a correlation between the age and number of treatments (linear model, adjusted R^2^ = 0.023, p-value = 0.00069). We also compared the parasite density (from microscopy done at the clinic during the 2011 season) with both molFOI for the 2011 season (c) and participant age (d). We found a significant negative correlation between participant age and density, as expected (linear model, adjusted R^2^ = 0.043, p-value = 1.3e^-6^), but we found no differences in the density data when grouped by participant molFOI in 2011 (Kruskal-Wallis test, □^2^(2) = 4.68, p-value = 0.096). Finally, we compared the parasite density data from the 2012-2016 samples with the molFOI per season (e) and the participants’ age per season (f). Here, we found a slight difference in density between the low and high molFOI groups (Kruskal-Wallis test, □^2^(2) = 10.52, p-value = 0.0052; Dunn test with Holm correction, p-values = 0.0037 (low vs. high), 0.17 (low vs. mid), and 0.11 (mid vs. high), but we did not find a significant relationship between age and parasite density (linear model, p-value = 0.14). Violin plots represent the density of the distribution, with horizontal lines at the 25th, 50th, and 75th percentiles. Parasite densities (c-f) are plotted on log scales. Abbreviations: molFOI, molecular force of infection.

**Extended Data Figure 3.**
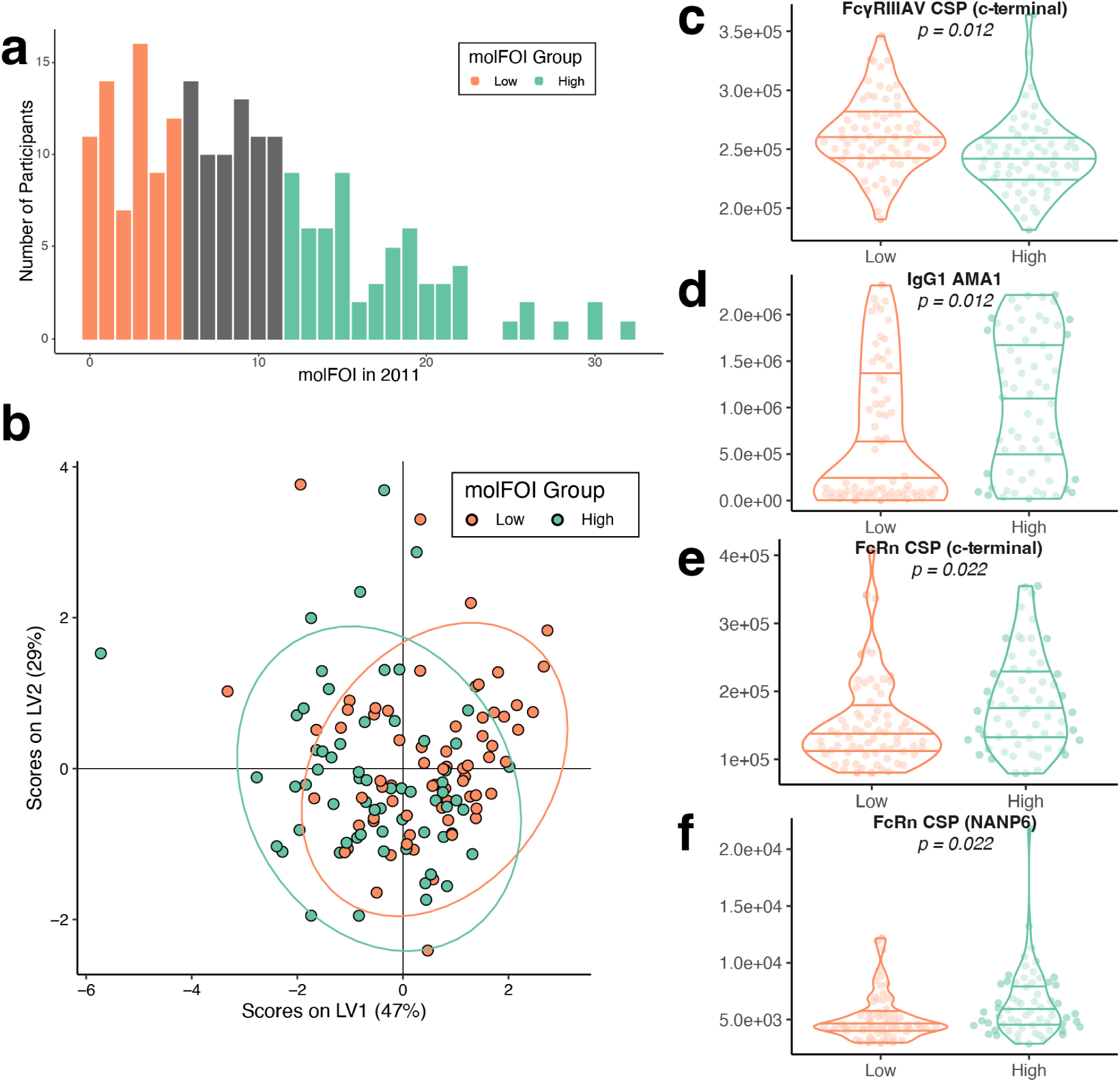
Functional antibody features distinguish between the top and bottom thirds of the molFOI distribution. The least stringent set of cutoffs that we used to determine the low and high molFOI groups. (a) Distribution of molFOI in the 2011 season for the 201 participants for whom we had both genetic and serology data. We compared low and high molFOI participants in the following analyses, who are colored in orange and teal, respectively. (See **Extended Data Table 1** for details on molFOI ranges used here and in other versions of this analysis). (b) We used the LASSO algorithm to select features that stratified the low and high molFOI groups. This plot shows the partial least squares latent variable (LV) scores based on the selected features. (c-f) Raw data from the 4 selected features that were selected by the LASSO algorithm. Each point represents a single participant. p-values are from Mann-Whitney tests, with Benjamini-Hochberg corrections. Violin plots represent the density of the distribution, with horizontal lines representing the 25th, 50th, and 75th percentiles. Abbreviations: AMA1, apical membrane antigen; IgG, immunoglobulin G; LV, latent variable; molFOI, molecular force of infection; NANP6, repeat region within circumsporozoite protein.

**Extended Data Figure 4.**
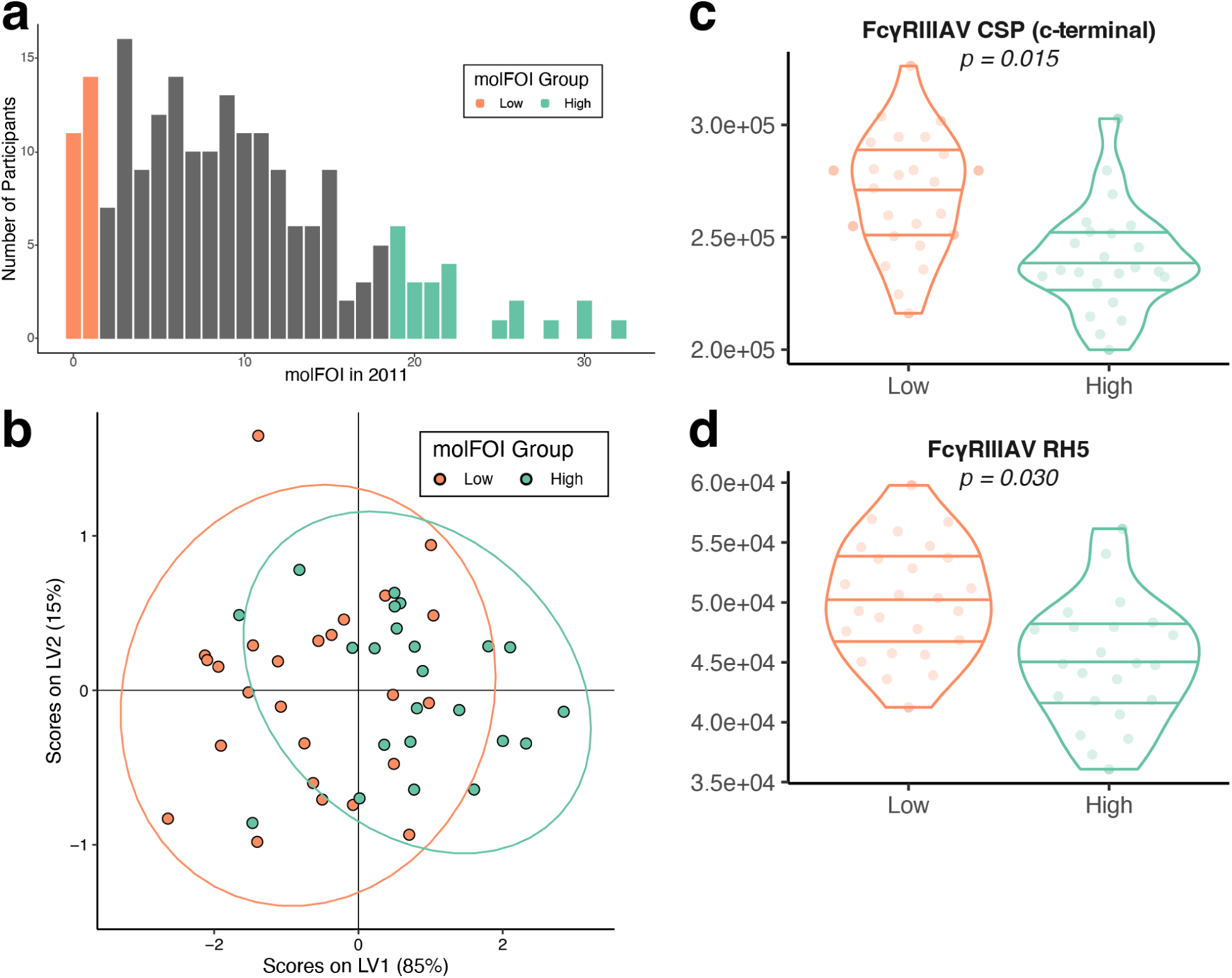
Immunological features distinguish between the top and bottom 12% of the molFOI distribution. The most stringent set of cutoffs that we used to determine the low and high molFOI groups. (a) Distribution of molFOI in the 2011 season for the 201 participants for whom we had both genetic and serology data. We compared low and high molFOI participants in the following analyses, who are colored in orange and teal, respectively. (See **Extended Data Table 1** for details on molFOI ranges used here and in other versions of this analysis). (b) We used the LASSO algorithm to select features that stratified the low and high molFOI groups. This plot shows the partial least squares latent variable (LV) scores based on the selected features. (c-d) Raw data from the 2 selected features that were selected by the LASSO algorithm. Each point represents a single participant. p-values are from Mann-Whitney tests, with Benjamini-Hochberg corrections. Violin plots represent the density of the distribution, with horizontal lines representing the 25th, 50th, and 75th percentiles. Abbreviations: LV, latent variable; molFOI, molecular force of infection; RH5, reticulocyte-binding protein homolog 5.

**Extended Data Figure 5.**
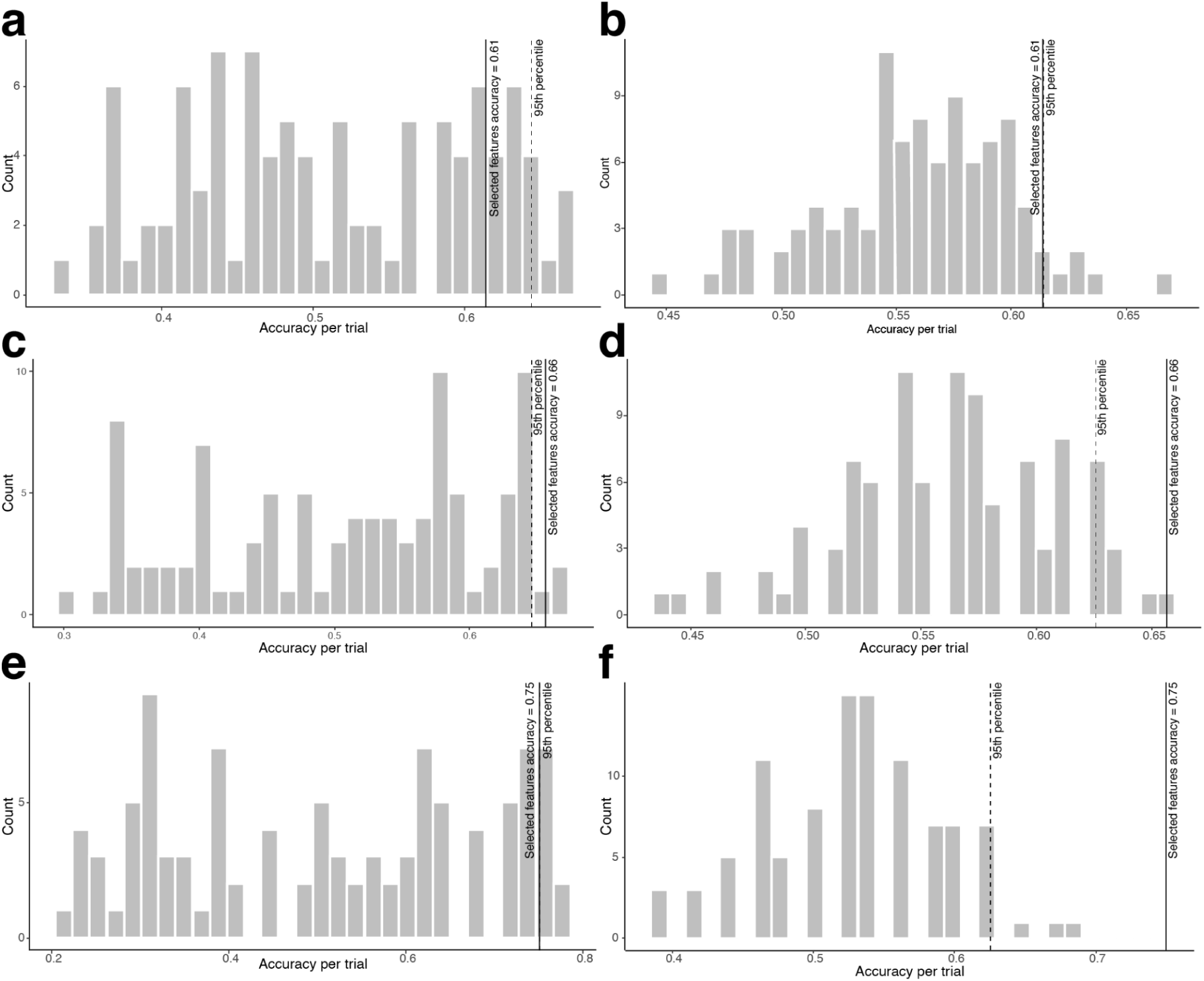
Cross-validation of PLS-DA classifiers. We used two types of negative control models to compare to the results from the PLS-DA classifiers we built from the systems serology data. (a,c,e) One set of models involved permuting the “low molFOI” and “high molFOI” group labels, then running the LASSO feature selection algorithm as before. (b,d,f) The other set of models involved keeping the group labels the same but selecting random features instead of going through the LASSO selection algorithm. In all panels, gray bars show the distribution of accuracy, from ten rounds of cross-validation, which included 10 permuted group trials and 10 random feature selection trials per round. The solid lines in each panel show the accuracy from the experimental (not control) classifiers, and the dotted lines show the 95th percentile of the accuracy from the cross-validation trials. Panels (a-b) are from the least stringent molFOI cutoffs, shown in **Extended Data Figure 3**. Panels (c-d) are from the middle set of molFOI cutoffs, shown in Figure 6. Panels (e-f) are from the most stringent molFOI cutoffs, shown in **Extended Data Figure 4**. Abbreviations: molFOI, molecular force of infection.

